# Mapping intra-tumoural heterogeneity in a spectrum of adolescent central nervous system tumours using APT-CEST and ^18^F-choline PET-MRI

**DOI:** 10.64898/2025.12.22.25339845

**Authors:** Trung Nguyen, Marilena Rega, Francisco Torrealdea, Joe Hearle, Moritz Zaiss, Ananth Shankar, Xavier Golay, Harpreet Hyare

**Author notes:** These authors share first authorship (Trung Nguyen, Marilena Rega). These authors share last authorship (Xavier Golay, Harpreet Hyare). **Corresponding author:** Dr Harpreet Hyare, University College London Hospitals NHS Foundation Trust, London, UK.

## Abstract

**Background:** Paediatric and adolescent gliomas and glioneuronal tumours remain challenging to assess non-invasively. Amide proton transfer (APT) chemical exchange saturation transfer (CEST) MRI has shown promise in adult gliomas but has not been well studied in younger patients.

**Purpose:** To assess whether APT CEST signal can act as a non-invasive surrogate of tumour proliferation in adolescent CNS tumours by correlating it with ^18^F-choline PET uptake (SUV) as a proxy for membrane synthesis / proliferative activity.

**Methods:** Ten adolescent patients (14–19 yrs) with confirmed or suspected gliomas / glioneuronal tumours underwent simultaneous APT CEST and ^18^F-choline PET-MRI. Regions of interest (ROIs) corresponding to non-enhancing, enhancing, necrotic tumour, and contralateral white matter were delineated. Mean APT signal intensity (SI) and PET SUV were extracted per ROI. Nonparametric statistics and Spearman’s correlation analyses were performed.

**Results:** APT SI was significantly elevated in enhancing, non-enhancing, and necrotic tumour ROIs compared to normal white matter (p<0.001). ^18^F-choline SUV was elevated in enhancing and necrotic ROIs vs white matter, but not significantly so for non-enhancing tumour (p=0.02). A strong correlation between whole-tumour APT SI and ^18^F-choline SUV was seen (Spearman ρ=0.86, p<0.001).

**Conclusion:** Our results indicate that APT CEST is feasible in adolescents and may reflect proliferative tumour burden. The detection of elevated APT SI even in non-enhancing tumour regions suggests potential utility in monitoring non-contrast-enhancing disease. Larger cohorts and multimodal correlation (e.g. Ki-67, amino acid PET) are warranted to confirm and extend these findings.

## Introduction

Central nervous system (CNS) tumours are the leading cause of cancer death in children, and gliomas remain a major challenge in the adolescent population [Ostrom 2017; Georgakis 2017]. Imaging-based assessment of treatment response is impeded by pseudoprogression and pseudoresponse phenomena, limiting the reliability of conventional MRI (contrast enhancement, size) in many therapeutic contexts [Wen 2023]. Thus, there is a pressing need for advanced imaging biomarkers that go beyond morphology to probe tumour biology.

Chemical exchange saturation transfer (CEST) MRI techniques allow indirect imaging of exchangeable protons (e.g., amide protons) associated with endogenous mobile proteins and peptides, at higher spatial resolution than conventional spectroscopy [Wu 2016]. APT CEST exploits the exchange between amide protons (resonating ~3.5 ppm) and bulk water protons. Because malignant tissues tend to harbour elevated concentrations of mobile proteins and peptides—and potentially altered pH / microenvironment—APT SI may serve as a marker of tumour aggressiveness [Cai 2017; Tagao 2014; Bai 2017]. In adults, APT SI has been correlated with histologic grade, molecular markers (e.g. IDH mutation / MGMT methylation), and survival outcomes [Jiang 2023; Joo 2019, Yao 2019] in addition to differentiating tumour progression from pseudoprogression [Karimian-Jazi 2025; Essed 2024]. However, whilst there are some studies showing value in differentiating paediatric tumour subtypes [Rajendran et al., 2023], the general adoption in paediatric and adolescent tumour imaging is sparse.

We hypothesised that APT CEST SI might serve as a non-invasive biomarker of proliferation in adolescent CNS tumours, and that correlation with ^18^F-choline PET uptake (a marker of membrane turnover and proliferation) would provide preliminary validation. Choline is a precursor of one of the main phospholipids used in cell membranes and its uptake in cancer cells reflects cellular proliferation and rapid cell membrane metabolism through its incorporation in choline-transporting transmembrane systems. An elevation of phosphocholine and total choline-containing compounds is thus one of the most widely established characteristics of cancer cells [Ackerstaff 2003]. Choline can be radiolabelled and an increased PET SUV has been shown to correlate with overexpression of choline kinase in tumour cell membranes [Li 2013, Moreau 2019]. Although amino acid PET (e.g. ^11^C-MET, ^18^F-FET) is increasingly the clinical standard in glioma imaging [Wen 2023; Langen 2023; Henssen 2025], ^18^F-choline provides favourable spatial resolution and contrast for this proof-of-concept study [Garcia-Vicentes 2019].

Recent reviews of CEST in gliomas emphasize that APT signal is sensitive not only to protein concentration, but also to pH, magnetization transfer (MT) background, and saturation settings. Multi-pool modelling (e.g. Lorentzian fitting / REX metrics) may help disentangle competing contributions [Cronin 2024; Kroh 2023]. A recent review highlights that higher APTw signals tend to correlate with higher-grade gliomas, and that molecular phenotypes (e.g. IDH status) may modulate these signals in adult cohorts [Deng et al., 2024].

Here, we present a feasibility study of concurrent APT CEST and ^18^F-choline PET imaging in adolescents with CNS tumours, exploring the relationship between APT SI and PET uptake, and the ability of APT to characterize enhancing, non-enhancing, and necrotic tumour compartments.

### Patients and Methods

#### Patients

10 patients (male n=6, mean age 16.4 years, age range in decade 10-20 years) with either a confirmed glioma (n=7) on histopathology, or a suspected glioma based on conventional MRI imaging (n=3) were prospectively recruited for ^18^F-choline PET and APT CEST between April 2016 and September 2019. Patient characteristics including histopathology and molecular genetics, treatment received, and clinical outcomes are summarised in Table 1. In all instances, APT CEST and ^18^F-choline PET were performed simultaneously, thereby ensuring a good registration of images and accurate quantification as both scans were performed at the same time point.

**Table 1.** Main characteristics of patients.

| Patient | Sex | Clinical Presentation | Histopathology | WHO grade | Molecular Genetics | Treatment | Patient Outcomes |
| --- | --- | --- | --- | --- | --- | --- | --- |
| 1 | F | Headache, vomiting, visual deterioration | GBM | IV | *IDH* mutant<br>*ATRX* loss of expression<br>*H3* negative | Near total resection<br>Radiotherapy<br>Temozolomide | Alive, well and in continuous remission |
| 2 | F | Seizures | Diffuse astrocytoma | II | *IDH* mutant<br>*ATRX* loss of expression | Radiotherapy<br>Temozolomide | Alive, well and in continuous remission |
| 3 | F | Visual deterioration | Midline HGG | IV | *IDH* wildtype<br>*H3* negative | Radiotherapy<br>Temozolomide | Alive, well and in continuous remission |
| 4 | M | Seizures | Pilocytic astrocytoma | I | *IDH* wildtype<br>*ATRX* retained<br>*BRAF* mutation | Complete resection | Alive, well and stable disease |
| 5 | M | Papilloedema, visual deterioration, ataxia | Midline HGG brainstem | IV | *H3* mutant<br>*ATRX* retained | Radiotherapy<br>Temozolomide<br>PCV maintenance chemotherapy | Died of disease progression 15 months after diagnosis |
| 6 | M | Seizures | GBM | IV | Unknown | Re-irradiation | Died of disease progression 10 months after recurrence |
| 7 | M | Headache, weakness | Diffuse midline glioma | IV | *IDH* wildtype<br>*H3* mutant<br>*ATRX* loss of expression<br>*BRAF* negative | Radiotherapy<br>Temozolomide | Alive with disease, terminally-ill due to disease progression |
| 8 | F | Headache, seizures | Glioneuronal tumour | I | *IDH* wildtype<br>*ATRX* retained | Debulking with residual disease, decompression of ventricle | Alive, well and in continuous remission |
| 9 | M | Headache, papilloedema | Schwannoma | I | *IDH* wildtype | None required | Alive, well and in continuous remission |
| 10 | M | Headache, coordination difficulties, ataxia | Rosette-forming glioneuronal tumour | I | *BRAF* negative | Temozolomide and lomustine | Alive, well and stable disease |

Note: GBM = glioblastoma multiforme; HGG = high grade glioma; PCV = procarbazine, lomustine, vincristine

#### Imaging protocol

All PET studies were conducted on a Siemens mMR biograph with an eight-channel dedicated PET-MRI receive head coil. PET images were acquired 10 minutes following intravenous injection of ^18^F-choline with dose administered dependent on patient weight (3.5 MBq/kg up to a maximum of 250 MBq). Data were acquired for 30 minutes post injection (matrix size 344 x 344) and corrected for scatter (relative scatter correction) and a 2 mm full width at half maximum (FWHM) Gaussian filter was applied. Attenuation correction was performed in 3-dimensions (3 iterations, 21 subsets) using the standard μ-map generated by the ultra-short echo time (UTE) sequence on the PET-MRI. SUV images were calculated accounting for the time between injection and acquisition and the ^18^F-choline half-life.

PET data were acquired simultaneously with the MRI sequences under standard resting conditions. The MRI protocol included an axial T_2_-weighted (w) inversion-recovery acquisition (TE/TR 397/5000, slice thickness 4 mm), an axial T_2_w turbo-spin-echo (TSE) sequence (TE/TR 300/4000, 4 mm), and an axial T_1_w 3D isovolumetric sequence before and after gadolinium (0.2 ml/kg) injection (TE/TR 3.8/2000, voxel size 1.1 × 1.1 × 1.1 mm). Diffusion-weighted imaging (DWI) (TE/TR 100/8000, 5 mm) was acquired using a single-shot echo-planar sequence in the axial plane with three b-values (0, 500 and 1000 s/mm^2^). CEST was acquired at two B_1_ saturation powers (0.75 and 1. 25 μT equivalent power) to allow for B_1_ field homogeneity correction with a snapshot CEST acquisition [19]. Saturation was achieved with a 3 s pulse train (Gaussian shaped pulses, 50 ms duration each, 50% duty cycle). A full z-spectrum was acquired for each B_1_ power with 41 frequency offsets ranging between ±5 ppm, sampled in alternative mode from −5 ppm to 0 ppm. A reference scan was also acquired at the start of the sequence for normalisation purposes. Saturation was followed by a single-shot centric-spiral reordered 3D gradient echo readout with an 8° flip angle, TE = 1.57 ms, TR = 1.57 ms, GRAPPA factor 2 in the phase encode direction and a matrix size 108 x 112 x 8. The effective TR per frequency offset was 5.48 s (accounting for both the readout and the saturation times).

A WASAB1 scan was acquired and was used for field homogeneity corrections [20]. The WASAB1 preparation consisted of 1 rectangular pulse of 5 ms duration and B1 = 3.7 μT. A reference scan was acquired for normalisation purposes and for 31 frequency offsets ranging from −3 to 3 ppm in alternative mode. The readout used was the same as for the CEST acquisition and was followed by a 2 s delay (TR per frequency was 4.49 s). Z-spectra were corrected for B_0_ and B_1_ inhomogeneities and normalised at the reference. APT-weighted images were calculated as the Z-spectra asymmetry centred at 3.5 ppm (from 3.2 to 3.8 ppm), also known as the APT range.

### Image analysis

#### Qualitative

An experienced neuroradiologist (HH) assessed the conventional MRI scans using five criteria from the Visually AcceSAble Rembrandt Images (VASARI) MRI feature set schema (http://wiki.nci.nih.gov/display/CIP/VASARI): (1) tumour location, (2) size of tumour epicentre, (3) enhancement quality, (4) proportion enhancing of whole tumour, and (5) proportion necrosis. The maximal perpendicular diameters (mm) were also recorded. The ^18^F-choline SUV and APT-weighted images were scored independently as positive if there was focal uptake within the tumour that was greater than normal brain activity.

#### Quantitative

T_2_-FLAIR and T_1_w-post Gd images were used to draw regions of interest (ROI) of the whole tumour, including “non-enhancing”, “enhancing”, and “necrotic core” in the tumour, and healthy contralateral “white matter” using ITKsnap (http://www.itksnap.org/pmwiki/pmwiki.php). Areas with obvious artefact or signal from blood vessels were excluded from the ROI. The APT-weighted image, ^18^F-choline SUV map and FLAIR image were registered to the T_1_w-post contrast image. The ROIs were copied from the FLAIR and T_1_ w-post contrast images to the APT-weighted image and ^18^F-choline SUV map and the mean APT signal (APT_mean_) and mean SUV (SUV_mean_) were extracted for each of the ROI.

### Statistical analysis

All values are expressed as mean (standard deviation). The APT_mean_ and SUV_mean_ in the normal appearing white matter, enhancing, non-enhancing and necrotic core ROIs were compared using the Kruskal-Wallis test. Post-hoc tests were performed comparing each of the enhancing, non-enhancing and necrotic core ROIs to the corresponding normal appearing white matter ROI. Due to the number of calculations, the significance level was set at p<0.01. Correlations between APT_mean_ and SUV_mean_ in the whole tumour, enhancing, non-enhancing, and necrotic core were assessed using the Spearman’s correlation coefficient. Statistical analyses were performed using SPSS Statistics (IBM Version 19.0, Armonk, NY).

## Results

### Qualitative findings Conventional MRI

The appearances of the tumours on conventional MRI were variable. The majority of tumours (n=7) were supratentorial and unilateral in location, with three posterior fossa tumours that were central. The majority of tumours (n=8) demonstrated some degree of enhancement, with six demonstrating marked/avid enhancement and two demonstrating mild enhancement. Seven tumours demonstrated necrosis ranging from <5% to 33% necrosis, and all but two tumours demonstrated a non-enhancing component. Examples of conventional imaging for a range of patients are demonstrated in Figure 1. The conventional imaging features for all patients are summarised in Table 2 according to the VASARI MRI feature set.

**Table 2.** Conventional MRI, 18F-choline SUV and APT-weighted SI findings.

| Patient | 18F-Cho<br>PET SUV | APT-weighted<br>SI | VASARI MRI features |  |  |  |  |  |  |
| --- | --- | --- | --- | --- | --- | --- | --- | --- | --- |
|  |  |  | F1 Location | F2 Side | F4 Enhancement | F5 %<br>Enhancement | F7 % Necrosis | Longest axis<br>(mm) | Shortest axis<br>(mm) |
| 1 | + | + | Temporal | R | Marked/avid | 67-95 | 6-33 | 32.8 | 14.8 |
| 2 | - | + | Frontal | R | None | N/A | 0 | 33.1 | 21.9 |
| 3 | - | + | Pons | C | None | N/A | 0 | 28.8 | 28.4 |
| 4 | + | + | Temporal | L | Marked/avid | 6-33 | <5 | 58.8 | 35.7 |
| 5 | + | + | Pons | C | Minimal/mild | 6-33 | 6-33 | 49.0 | 34.8 |
| 6 | + | + | Frontal | L | Marked/avid | 6-33 | 6-33 | 142.9 | 69.4 |
| 7 | + | + | Thalamus | L | Minimal/<br>mild | <5 | 0 | 11.9 | 8.4 |
| 8 | + | + | Parietal | L | Marked/avid | 34-66 | <5 | 27.1 | 21.2 |
| 9 | + | + | Frontal | R | Marked/avid | >95 | <5 | 50.1 | 30.4 |
| 10 | + | + | Cerebellum | C | Marked/avid | >95 | <5 | 35.8 | 20.6 |

Note: + = increased signal in the tumour compared to normal brain; - = no increased signal in the tumour compared to normal brain; R = right; L = left; C = central

**Fig. 1.**
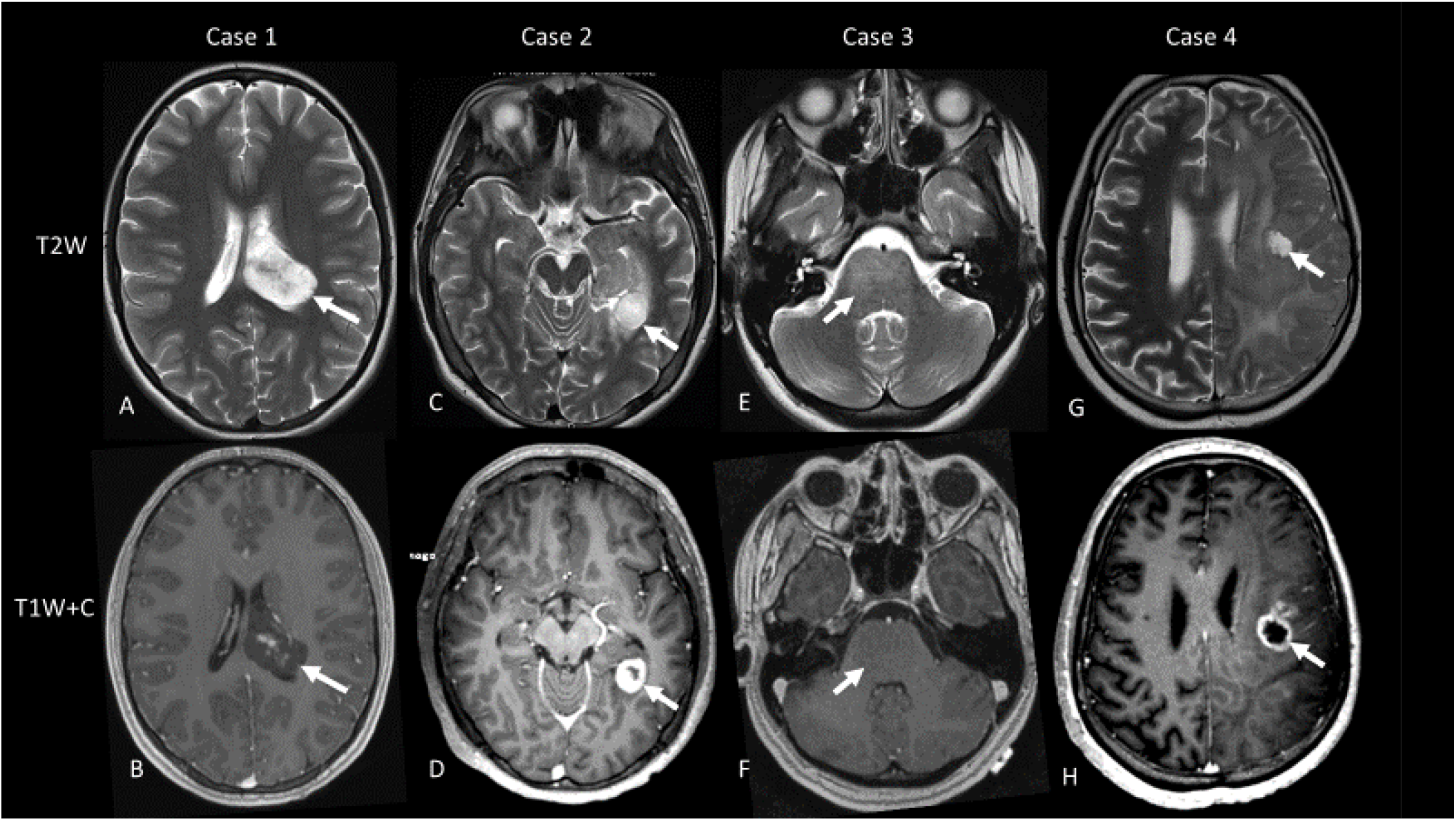
Spectrum of conventional MRI images in four patients. Case 1: glioneuronal tumour (WHO I) – T2w image demonstrates a large, well-defined tumour arising from the left peritrigonal white matter extending into the left trigone (A) with minimal enhancement (B). Case 2: pilocytic astrocytoma (WHO I) – T2w image demonstrates a left temporal periventricular signal abnormality (C) associated with ring enhancement post contrast (D). Case 3: midline high grade glioma (WHO IV, histone mutant) – diffuse signal abnormality is noted in the pons on the T2w image (E) with no pathological enhancement post contrast (F). Case 4: GBM (WHO IV) – T2w image demonstrates a large left hemispheric infiltrating lesion causing mass effect on the left lateral ventricle (G) associated with focal ring enhancement on the T1w image post contrast (H).

### ^18^F-choline PET-MRI and APT CEST

The majority of tumours (n=8) demonstrated ^18^F-choline PET avidity whilst all tumours demonstrated increased signal within the tumour compared to normal brain on the APT-weighted image. The two tumours that were ^18^F-choline PET SUV negative were also non-enhancing tumours. Examples of the SUV and APT-weighted maps obtained in three of the patients are demonstrated in Figure 2.

**Fig. 2.**
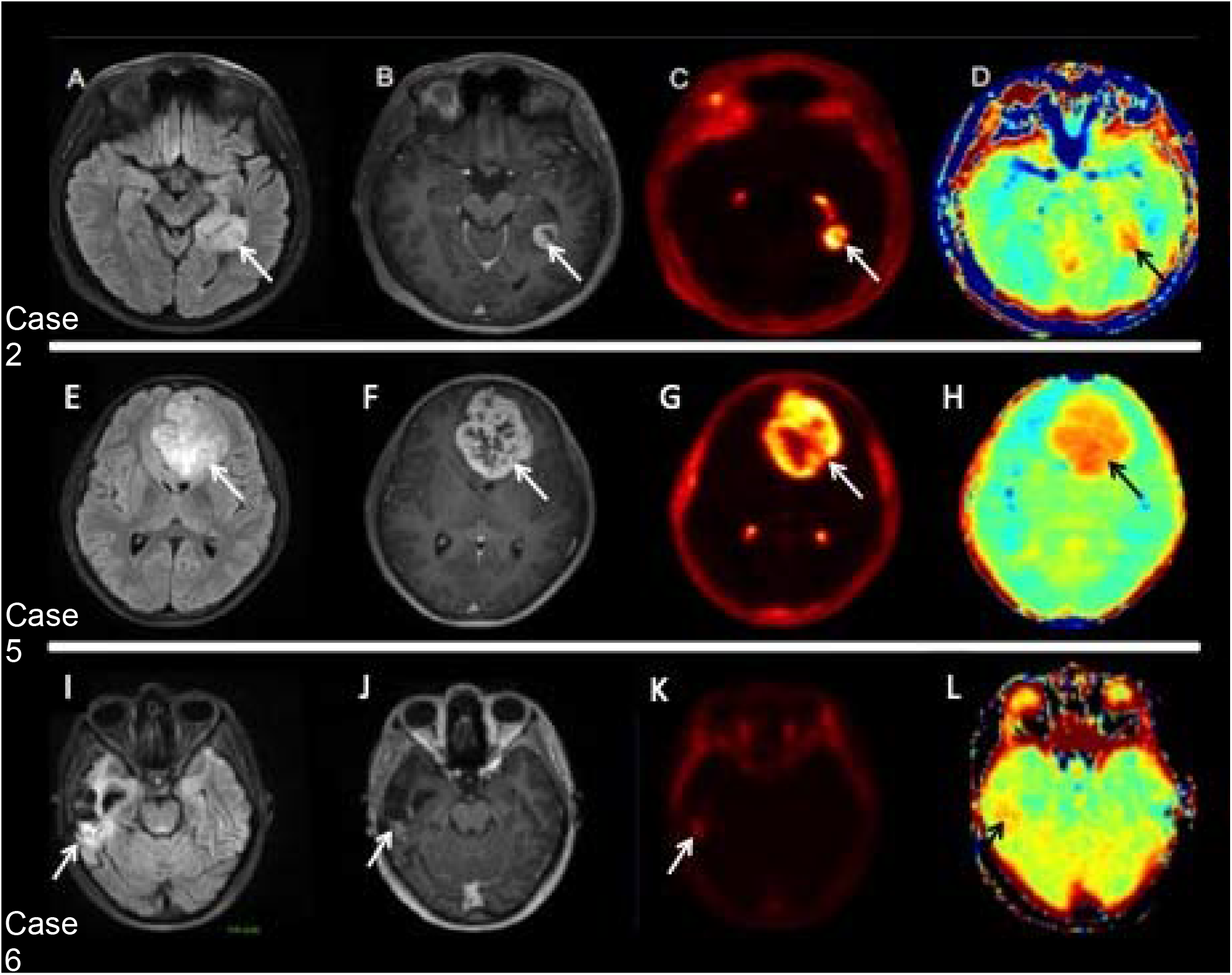
Examples of conventional MRI, 18F-choline SUV and APT-weighted maps in three patients. Case 2 (A-D) pilocytic astrocytoma (WHO I) – FLAIR demonstrates left temporal signal abnormality associated with ring enhancement on the T1w image post contrast, increased 18F-choline SUV and increased APT signal in the enhancing component. Case 5 (E – H) schwannoma (WHO I). FLAIR demonstrates a large midline lesion associated with avid heterogenous enhancement, increased 18F-choline SUV and increased APT signal throughout the tumour. Case 6 (I-L) GBM (WHO IV). FLAIR demonstrates the right temporal surgical bed with increased signal abnormality at the posterior margin (associated with mild nodular enhancement, focal increased 18F-choline SUV and focal increased APT signal within the enhancing component.

### Quantitative findings

#### Tumour ROI analysis

The overall means and standard deviations of the APT signal and ^18^F-choline PET SUV in each ROI for the patients are summarised in Table 3. The highest APT signal was seen in the enhancing and necrotic core ROI and the lowest APT signal was seen in the non-enhancing tumour ROI (Figure 3). The highest ^18^F-choline PET SUV was observed for the enhancing tumour ROI and the lowest was seen in the non-enhancing tumour ROI. There was a significant difference in APT signal for the enhancing, non-enhancing and necrotic ROI when compared to normal appearing white matter (p<0.001). Although, there was a significant difference in ^18^F-choline PET SUV for the enhancing and necrotic ROI compared to normal appearing white matter (p<0.001), there was no significant difference in ^18^F-choline PET SUV for the non-enhancing ROI versus normal appearing white matter (Figure 3, p=0.02).

**Table 3.** Summary of quantitative findings in tumour ROI.

| ROI | APT [a.u.] | 18F-cho PET [SUV] |
| --- | --- | --- |
| Enhancing<sup>a</sup> | 0.012 (0.004) | 1.281 (0.528) |
| Non-enhancing<sup>b</sup> | 0.002 (0.003) | 0.392 (0.368) |
| Necrotic core<sup>c</sup> | 0.019 (0.004) | 0.998 (0.422) |
| Normal appearing white matter<sup>d</sup> | -0.009 (0.004) | 0.186 (0.057) |
| p value a vs d | <0.001 | <0.001 |
| p value b vs d | <0.001 | 0.02 |
| p value c vs d | <0.001 | <0.001 |

Note: All values are expressed as mean (standard deviation). APT = amide proton transfer; a.u. = arbitrary units; 18F-cho = 18F-fluoromethylcholine; SUV = standardised uptake value

**Fig. 3.**
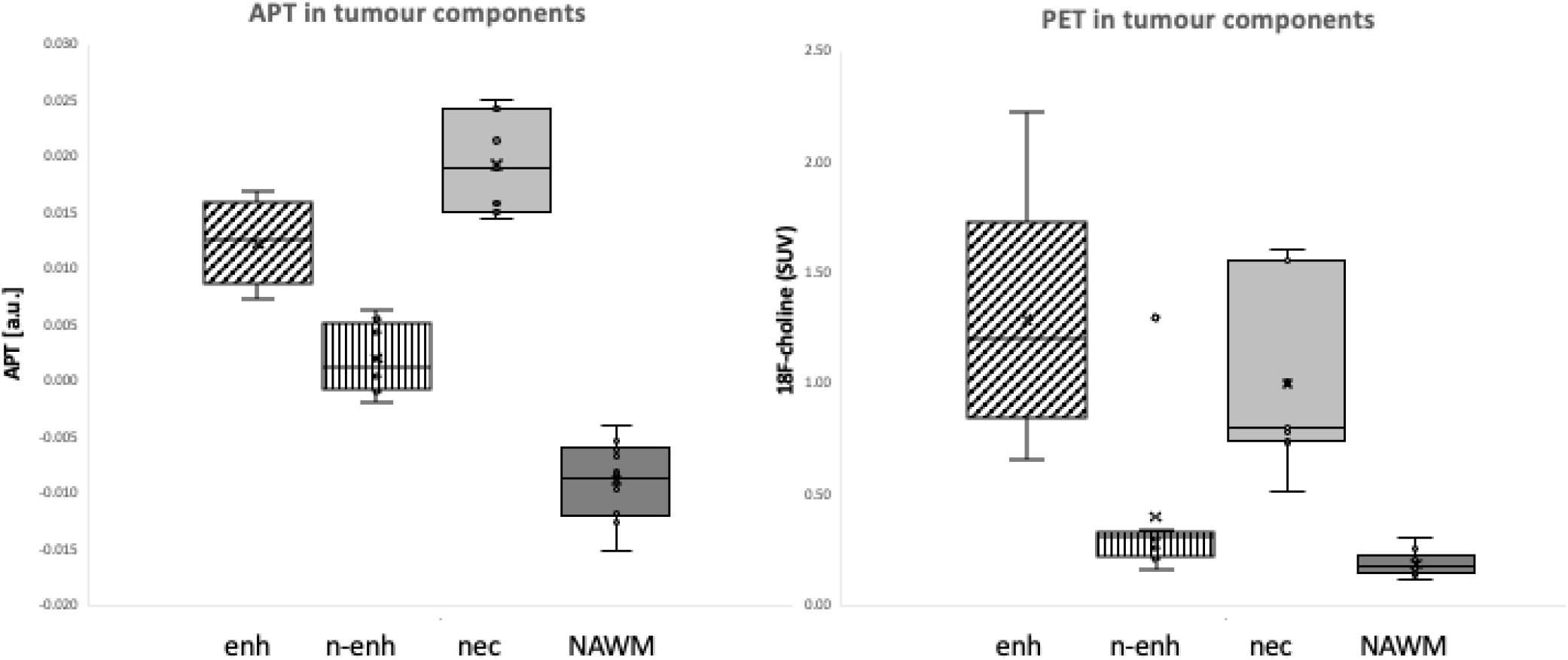
APT signal and 18F-choline PET SUV in enhancing, non-enhancing, necrotic and healthy appearing white matter ROI. Box plot demonstrating APT signal and ^18^F-choline PET SUV in the enhancing, non-enhancing, necrotic core and normal appearing white matter ROI. Note: enh = enhancing ROI; n-enh = non-enhancing ROI; nec = necrotic core ROI; NAWM = normal appearing white matter ROI; APT = amide proton transfer; a.u. = arbitrary units; ^18^F-choline = fluoromethylcholine; SUV = standardised uptake value.

#### Correlation of APT signal and^18^ F-choline PET SUV

The correlation between the different metrics was also assessed using the Spearman’s correlation coefficient. A strong correlation was observed for whole tumour APT signal when compared to whole tumour ^18^F-choline PET SUV (Spearman ρ=0.86, p<0.001). A weaker correlation was seen for APT signal versus ^18^F-choline PET SUV for the enhancing tumour ROI (Spearman ρ=0.63, p<0.001) and no correlations were found between APT signal and ^18^F-choline PET SUV for the non-enhancing and necrotic tumour ROI. Figure 4 demonstrates the relationship between APT signal and ^18^F-choline PET SUV in the whole tumour and enhancing tumour ROIs.

**Fig. 4.**
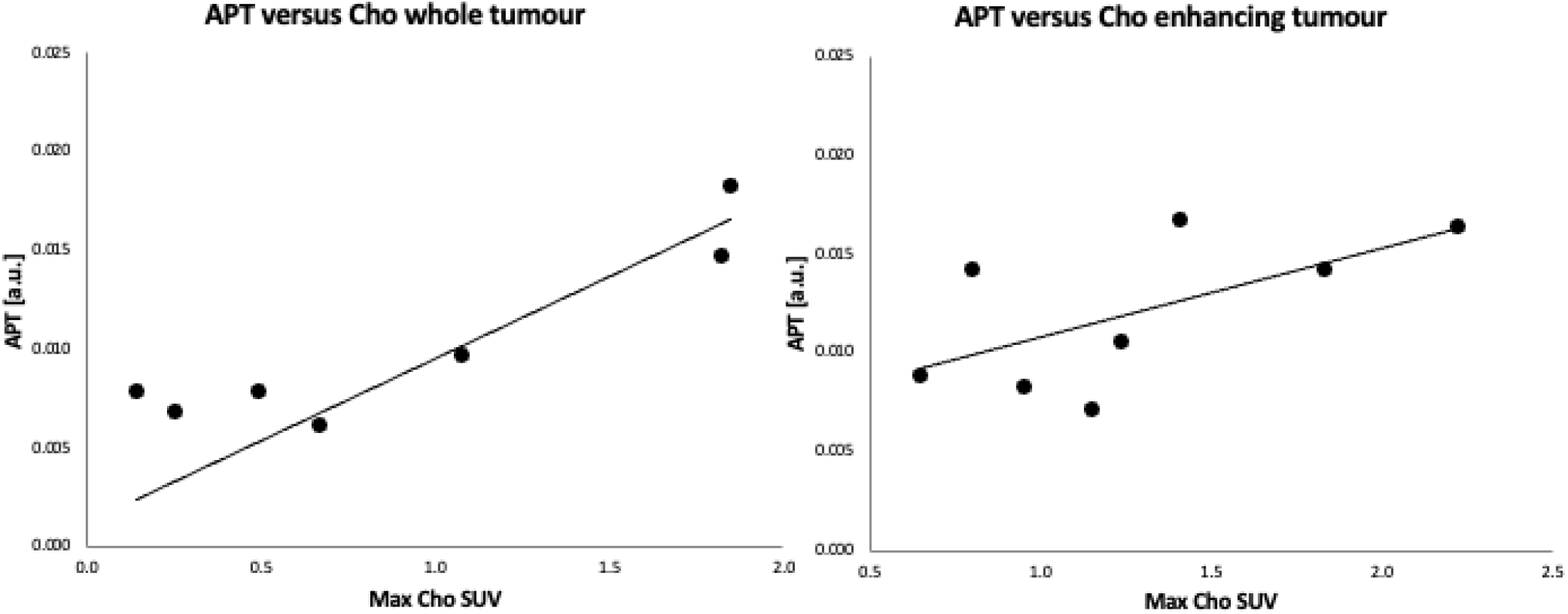
Relationship between APT-CEST and ^18^F choline SUV in whole tumour and enhancing tumour ROIs. Relationship between APT signal and ^18^F-choline PET SUV in the whole tumour and enhancing ROIs: whole tumour 18F-choline PET SUV (Spearman *ρ*=0.86, p<0.001), enhancing tumour ROI (Spearman *ρ*=0.63, p<0.001). Note: enh = enhancing ROI; n-enh = non-enhancing ROI; nec = necrotic core ROI; NAWM = normal appearing white matter ROI; APT = amide proton transfer; a.u. = arbitrary units; Cho = fluoromethylcholine; SUV = standardised uptake value.

## Discussion

We demonstrate the technical feasibility of APT CEST in paediatric and adolescent CNS tumours and demonstrate that APT SI can map intra-tumoural heterogeneity. We demonstrate a strong correlation with ^18^F-choline PET uptake at the whole-tumour level, suggesting that APT-CEST is a sensitive biomarker of tumour proliferation. Importantly, elevated APT SI was demonstrated in non-enhancing tumour regions suggests potential sensitivity to non-contrast-enhancing infiltrative disease.

In adult glioma cohorts, APT/CEST has been explored extensively. CEST was first used in rat models to differentiate normal brain tissue from glioma tissue [Cai 2015], predict histological grading of gliomas and to assess therapeutic response to temozolomide in murine models [Zhou 2017]. Studies in human adults also suggest CEST might be useful for predicting histological grade of gliomas [Tagao 2014, Bai 2017, Harris 2016], IDH mutation status/MGMT promoter methylation status [Paech 2018, Joo 2019], assessing response to treatment [Yao 2019; Regnery 2018] and be an independent predictor of poor overall survival and progression-free survival in high-grade glioma [Paech 2019].

A major challenge for clinical adoption is reproducibility. A recent multi-scanner / multi-session reproducibility study (19 healthy volunteers, 7 brain tumour patients) at 3T reported low coefficients of variation (COV) for Lorentzian-difference (LD) and relaxation-compensated metrics (MTR_Rex) (within-session WM COV ~0.56%, between sessions ~0.84%) and somewhat higher variability in tumour ROIs (~4–6 %) but acceptable stability across sessions and scanners [Wu et al., 2023]. This suggests that advanced fitting methods (beyond simple MTR_asym) may offer more robust quantification, particularly for longitudinal tracking. Another study [Kroh et al. 2023] evaluated semi-solid MT and APT-based metrics (asymmetry, Lorentzian fits) to predict clinical endpoints and argued that combining CEST and MT modelling helps isolate biologically meaningful contrast while suppressing confounds (MT, spillover).

Recently, APT CEST has also been applied to the problem of distinguishing glioma recurrence from pseudoprogression. A 2025 study in recurrent glioma patients (30 progression, 12 pseudoprogression) showed that APTw values were significantly higher in progressing disease than in pseudoprogression (1.99% vs 1.26%, p = 0.008). Additionally, APT revealed focal “metabolic hotspots” within FLAIR lesions not evident on conventional MRI, and early changes in resection cavity APT even before contrast changes [Karimian-Jazi et al., 2025]. This supports the potential of APT for treatment monitoring beyond static imaging.

In paediatric neuro-oncology, APT CEST remains nascent. A brief initial experience paper outlines technical challenges (motion, B0/B1 inhomogeneity, smaller brain size) and potential value, but few case series exist. Meanwhile, reviews of advanced neuroimaging in paediatric brain tumours list CEST and molecular MRI among the promising emerging modalities [Nikam et al., 2022].

To our knowledge, this is the first study to pair APT CEST with ^18^F-choline PET uptake in adolescent CNS tumours. The strong correlation at the whole-tumour level supports the hypothesis that APT SI is sensitive to tumour proliferation / metabolic turnover. Increased ^11^C-choline (Cho) PET SUV has been shown to correlate with cell proliferation indices in lung cancer biopsy specimens thought to be due to overexpression of choline kinase in cancer cell membranes [Li 2013]. Several previous studies have evaluated the role of Cho PET/CT in low- and high-grade gliomas [Gomez-Rio 2015, Fraioli 2022] and the good tumour to normal brain contrast and the high spatial resolution also influenced our choice. The absence of correlation in necrotic or non-enhancing subregions may reflect tissue heterogeneity, limited PET sensitivity in diffuse regions, or confounding contributions (e.g. extracellular proteins, pH effects, MT background).

The observation of high APT SI in necrotic ROIs is intriguing and may reflect accumulation of breakdown products (mobile peptides/proteins) or altered water environment. The lack of correlation with PET in necrosis supports a non-proliferative origin of signal. APT SI is influenced not only by mobile protein / peptide concentration, but also pH, water content, exchange rate kinetics, magnetization transfer background, and direct saturation effects. Multi-pool modelling (LD fitting, MTR_Rex) may help isolate the amide exchange contribution more reliably [Kroh et al., 2023].

The non-enhancing tumour ROI showed elevated APT versus white matter and is supported by reports in the adult population including a study of 18 patients where the heterogeneity of non-enhancing gliomas was mapped using APT-weighted signal [Wagnert 2021]. However, PET uptake did not differ significantly. This may stem from limited sensitivity of choline PET in diffuse infiltrative disease, partial volume effects, or mismatch between proliferation and choline metabolism in lower-density regions. Indeed, image-targeted biopsies to correlate with cell density would strengthen claims of APT as a proliferative biomarker. Alternatively, amino-acid PET tracers (^11^C-MET, ^18^F-FET and ^18^F-DOPA) that have been shown to be more specific for glioma metabolic activity [Soni 2023], may provide improved correlation with CEST signals. Whilst a single study of 8 patients comparing APT CEST with ^18^F-FET demonstrated no correlation between the two techniques [da Silva 2018], and another showed higher diagnostic performance for APT compared to methionine for post-treatment gliomas [Park 2018], future larger simultaneous APT / amino-acid PET studies could provide more direct biological linkage [Langen et al., 2023].

In summary, our results provide promising evidence that APT CEST is technically feasible in paediatric and adolescents brain tumours and that APT SI correlates with PET-derived proliferation metrics at the global tumour level. The sensitivity of APT to non-enhancing tumour components offers an attractive supplement to traditional imaging, particularly in diffuse gliomas. Future work will expand cohorts, validate histologically, and explore longitudinal and multimodal integration.

## Conclusion

This study presents the first simultaneous correlation of APT CEST and ^18^F-choline PET in adolescent CNS tumours, demonstrating feasibility, compartmental discrimination, and a strong imaging–metabolic correlation. We propose that APT CEST may serve as a surrogate biomarker of tumour proliferation and could aid in assessing non-enhancing disease activity. Further prospective studies integrating amino acid PET, histology, and longitudinal follow-up are warranted to validate and refine the role of APT CEST in paediatric neuro-oncology.

## Data Availability

All data produced in the present study are available upon reasonable request to the authors.

## Statements and Declarations

This work was supported by funding from the European Union’s Horizon 2020 programme (Grant No. 667510) and the NIHR UCLH BRC. The authors have no relevant financial or non-financial interests to disclose. All authors contributed to study design, data acquisition, analysis, and manuscript drafting. The first draft was written by Trung Nguyen and Marilena Rega. All authors read and approved the final manuscript. Ethics approval was granted by the UCL/UCLH Research Ethics Committee (REC 07/Q0502/15). Written informed consent was obtained from all participants following institutional board approval.

